# An artificial neural network approach integrating plasma proteomics and genetic data identifies *PLXNA4* as a new susceptibility locus for pulmonary embolism

**DOI:** 10.1101/2020.10.05.20207001

**Authors:** Misbah Razzaq, Maria Jesus Iglesias, Manal Ibrahim-Kosta, Louisa Goumidi, Omar Soukarieh, Carole Proust, Maguelonne Roux, Pierre Suchon, Anne Boland, Delphine Daiain, Robert Olaso, Lynn Butler, Jean-François Deleuze, Jacob Odeberg, Pierre-Emmanuel Morange, David-Alexandre Trégouët

## Abstract

Venous thromboembolism is the third common cardiovascular disease and is composed of two entities, deep vein thrombosis (DVT) and its fatal form, pulmonary embolism (PE). While PE is observed in ∼40% of patients with documented DVT, there is limited biomarkers that can help identifying patients at high PE risk.

To fill this need, we implemented a two hidden-layers artificial neural networks (ANN) on 376 antibodies and 19 biological traits measured in the plasma of 1388 DVT patients, with or without PE, of the MARTHA study. We used the LIME algorithm to obtain a linear approximate of the resulting ANN prediction model. As MARTHA patients were typed for genotyping DNA arrays, a genome wide association study (GWAS) was conducted on the LIME estimate. Detected single nucleotide polymorphisms (SNPs) were tested for association with PE risk in MARTHA. Main findings were replicated in the EOVT study composed of 143 PE patients and 196 DVT only patients.

The derived ANN model for PE achieved an accuracy of 0.89 and 0.79 in our training and testing sets, respectively. A GWAS on the LIME approximate identified a strong statistical association peak (p = 5.3×10^−7^) at the *PLXNA4* locus, with lead SNP rs1424597 at which the minor A allele was further shown to associate with an increased risk of PE (OR = 1.49 [1.12 – 1.98], p = 6.1×10^−3^). Further association analysis in EOVT revealed that, in the combined MARTHA and EOVT samples, the rs1424597-A allele was associated with increased PE risk (OR = 1.74 [1.27 – 2.38, p = 5.42×10^−4^) in patients over 37 years of age but not in younger patients (OR = 0.96 [0.65 – 1.41], p = 0.848).

Using an original integrated proteomics and genetics strategy, we identified *PLXNA4* as a new susceptibility gene for PE whose exact role now needs to be further elucidated.

**Author Summary:** Pulmonary embolism is a severe and potentially fatal condition characterized by the presence of a blood clot (or thrombus) in the pulmonary artery. Pulmonary embolism is often the consequence of the migration of a thrombus from a deep vein to the lung. Together with deep vein thrombosis, pulmonary embolism forms the so-called venous thromboembolism, the third most common cardiovascular disease, and its prevalence strongly increases with age. While pulmonary embolism is observed in ∼40% of patients with deep vein thrombosis, there is currenly limited biomarkers that can help predicting which patients with deep vein thrombosis are at risk of pulmonary embolism. We here deployed an Artificial Intelligence based methodology integrating both plasma proteomics and genetics data to identify novel biomarkers for PE. We thus identified the *PLXNA4* gene as a novel molecular player involved in the pathophysiology of pulmonary embolism. In particular, using two independent cohorts totalling 1,881 patients with venous thromboembolism among which 467 experienced pulmonary embolism, we identified a genetic polymorphism in the *PLXNA4* gene that associates with ∼2 fold increased risk of pulmonary embolism in patients aged more than ∼40 years.

## Introduction

Deep vein thrombosis (DVT) and Pulmonary Embolism (PE) are often considered as two sides of the same coin, venous thromboembolism (VTE), the third most common cardiovascular disease. VTE is a complex disease resulting from the interplay of various factors including (epi-)genetics and environmental sources. VTE incidence is estimated at 1 per 1000 patient-years, and its fatal form, PE, is associated with a mortality rate of 6% in the acute phase and 20% after one year [1]. PE generally results from the migration of a blood clot from a deep vein to the lung and is observed in ∼40% of patients with documented DVT [2]. However, isolated PE without any trace of DVT can also be observed either when the clot has completely migrated to the lung or when it is a pulmonary clot in situ as recently highlighted in COVID-19 patients [3]. Even though some specific risk factors for PE have been identified in DVT patients such as obesity, sickle cell disease [4] as well as some genetic variations in *F5* [4] and *GRK5* [5] genes, the exact, likely multifactorial, biological mechanisms that lead to PE are still not fully characterized. Besides, there is still limited biomarkers that can help discriminating patients that will develop PE from those who won’t, the former being then at higher risk of death. Thus, there is clearly a need for novel PE-associated molecular markers to be identified.

Plasma is an ideal potential source for VTE biomarkers; the intravascular compartment itself is the site of disease manifestation and tests are relatively non-invasive, quick and cheap. Several types of molecular determinants can be assessed in plasma samples including microRNAs, metabolites and proteins, and all of them have been investigated in the context of VTE. For example, plasma microRNAs have been assessed in relation to VTE recurrence [6,7]. Plasma proteomics has been employed to discover novel proteins associated with VTE risk [8,9] and plasma metabolomics used to identify novel mechanisms involved in VTE etiology [10,11]. Only one study has so far adopted an exploratory plasma proteomics strategy to identify novel proteins associated with high-risk versus low-risk of PE in humans. This study [12] was based on a relatively small sample size and compared 6 patients with high risk of PE to 6 patients at low PE risk, risk being classified based on clinical presentations and symptoms, with plasma samples profiled by matrix-assisted laser desorption/ionization–time-of-flight/time-of-flight mass spectrometry (MALDI-TOF/TOF MS).

In this work, we aim at identifying novel molecular phenotypes that could help in better characterizing the biological mechanisms involved in the development of PE in VTE patients. For this, 234 plasma proteins targeted with 376 protein specific antibodies, with the major part derived from the Human Protein Atlas (HPA) repository [13] were profiled in 1388 VTE patients selected from the MARTHA study [14,15] and from whom 283 had experienced a symptomatic PE event. To explore far beyond the search for linear associations between protein levels and PE risk and to identify more complex relationships that could serve as integrative markers of upstream/downstream mechanisms involving molecular determinants that have not necessarily been measured, we deployed a sequential procedure implementing several methodologies selected from the deep-learning domain. Briefly, and as summarized in Figure 1 and more detailled thereafter, the first step consists in applying an under-sampling algorithm (edited nearest neighbors) [16] to remove individuals with strong data heterogeneity that would hamper the efficiency of the downstream analyses, leaving to subsample of 592 VTE patients (497 DVT and 95 PE). This subsample was then used in an Artifical Neural Network (ANN) learning framework in order to predict PE from proteomics data. We then used the Local Interpretable Model-agnostic Explanations (LIME) algorithm [17] to derive a linear approximate of the ANN based predictor for PE risk which would, in addition, have a more meaningful biological interpretation. As MARTHA patients have been previously typed for genome-wide genotype data, we then conducted a genome wide association study of the LIME predictor of PE in order to detect single nucleotide polymorphisms (SNPs) associated with the predictor with the hope that the integration of genetic and proteomic data could provide additional insights into the pathophysiology underlying the identified predictor [18,19]. SNPs with strong statistical association with the LIME predictor were tested for association with PE risk in the whole original MARTHA dataset and significant associations were further tested for replication in an independent study of 339 VTE patients including 143 with PE. Sequencing data were also scrutinized in some patients with observed VTE outcomes poorly predicted by our ANN/LIME prediction models in order to identify rare variants that could be responsible for the observed phenotypes.

**Figure 1.**
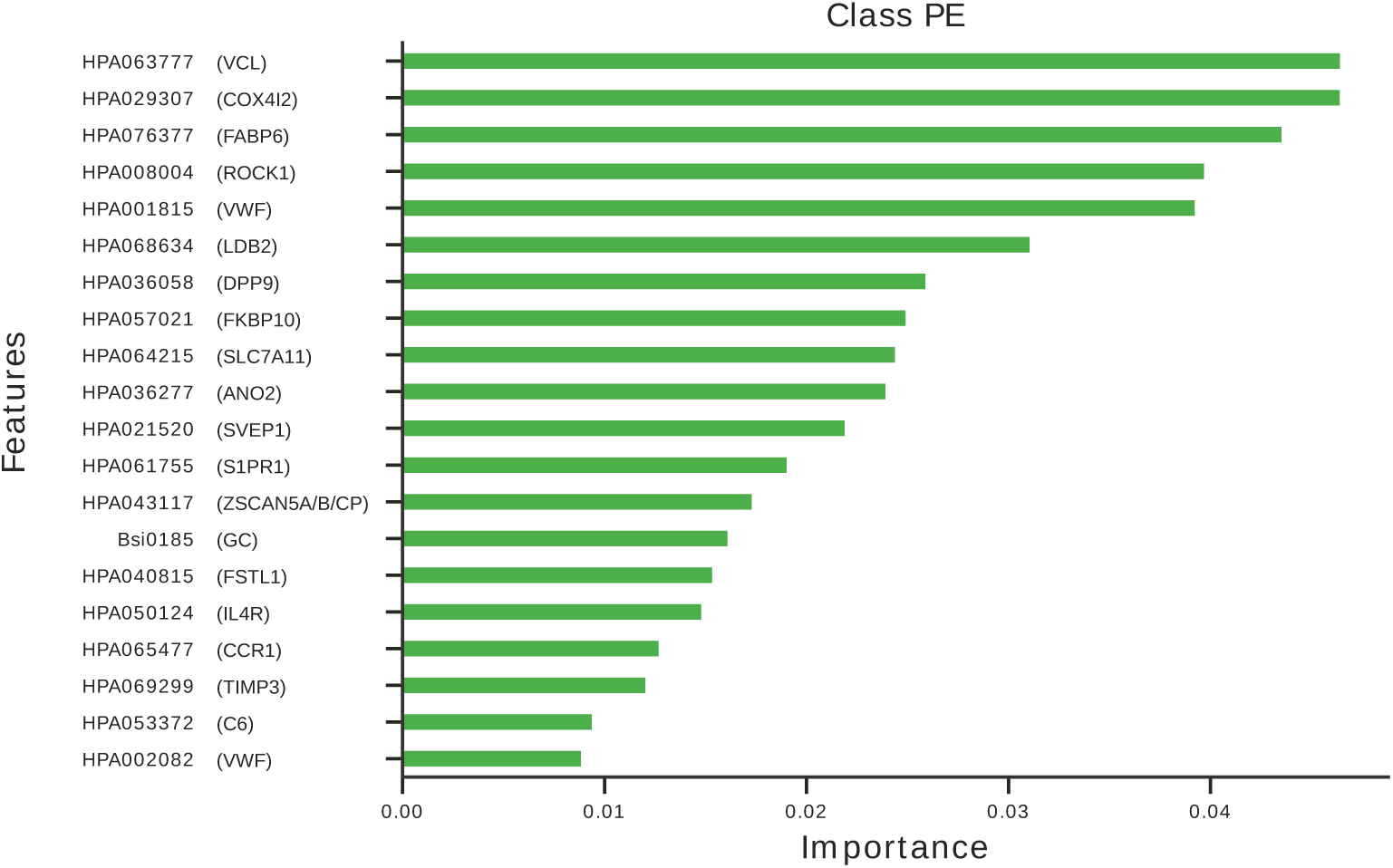
Analysis workflow of the present study

## Results

### Data description

The MARTHA proteomics substudy was composed of 1,388 VTE patients among which 1,105 were diagnosed for DVT, 95 with isolated PE and 188 with both DVT and PE (Table 1). Patients were phenotyped for 19 quantitative traits known to be involved in thrombotic biological processes (Supplementary Table 1) and for 234 different proteins using targeted affinity proteomics with 376 protein specific antibody reagents using multiplexed suspension bead array technology. These proteins were selected for 1) their known roles in the coagulation/fibrinolysis cascade and/or intermediate traits of relevance to thrombosis, 2) their specific expression in endothelial cells (a key cell type involved in thrombosis physiopathology) or 3) encoded by genes identified in pangenomic studies as associated with several cardiovascular disease-linked biological pathways (e.g platelet function, renal function, inflammation). The list of antibody reagents with their target proteins is given in Supplementary Table 2.

**Table 1.** Characteristics of the MARTHA proteomics study

|  | DVT | PE | DVT+PE |
| --- | --- | --- | --- |
| N | 1105 | 95 | 188 |
| Age at sampling | 46.67 (14.90) | 48.63 (15.26) | 51.57 (16.99) |
| Age at first VTE | 40.89 (15.28) | 41.64 (15.02) | 44.22 (17.56) |
| Female sex | 716 (65%) | 78 (82%) | 112 (60 %) |
| Women under oral contraceptives at VTE event | 286 (26%) | 35 (37%) | 45 (24%) |
| FV Leiden (rs6025) heterozygotes | 255 (23%) | 17 (18%) | 39 (21%) |
| Anticoagulant therapy at plasma sampling | 303 (27%) | 29 (31%) | 76 (40%) |
| Smokers | 209 (19 %) | 18 (19 %) | 24 (13 %) |
| BMI | 25.14 (4.57) | 25.20(4.39) | 26.43(4.62) |

DVT : Deep Vein Thrombosis ; PE : Pulmonary Embolism ; BMI : Body Mass Index Data shown correspond to mean (standard deviation) and count (percentage) for continuous categorical variables, respectively

**Table 2.** Individual predictions of VT event provided by ANN and LIME in the 16 patients of the testing set

| Individual | Observed clinical class | ANN prediction for class PE | Local Prediction for class PE |
| --- | --- | --- | --- |
| 1 | DVT | 0.04 | 0.31 |
| 2 | DVT | 0.00 | 0.18 |
| 3 | DVT | 0.03 | 0.24 |
| 4 | DVT | 0.02 | 0.17 |
| 5 | DVT | 0.00 | 0.23 |
| 6 | DVT | 0.02 | 0.32 |
| 7 | DVT | 0.00 | 0.25 |
| 8 | DVT | 0.04 | 0.22 |
| 9 | DVT | 0.25 | 0.34 |
| 10 | DVT | 0.88 | 0.26 |
| 11 | PE | 0.00 | 0.30 |
| 12 | PE | 0.20 | 0.31 |
| 13 | PE | 0.98 | 0.94 |
| 14 | PE | 1.0 | 1.0 |
| 15 | PE | 0.01 | 0.15 |
| 16 | PE | 0.80 | 0.77 |

Exploration of this dataset using high-dimensional visualization techniques including principal component analysis, t-SNE [20] and UMAP [21] did not reveal any specific stratification in the data nor outliers (Figure 2) but rather illustrates that the three class of patients (DVT, PE, DVT+PE) could not be easily separated.

**Figure 2.**
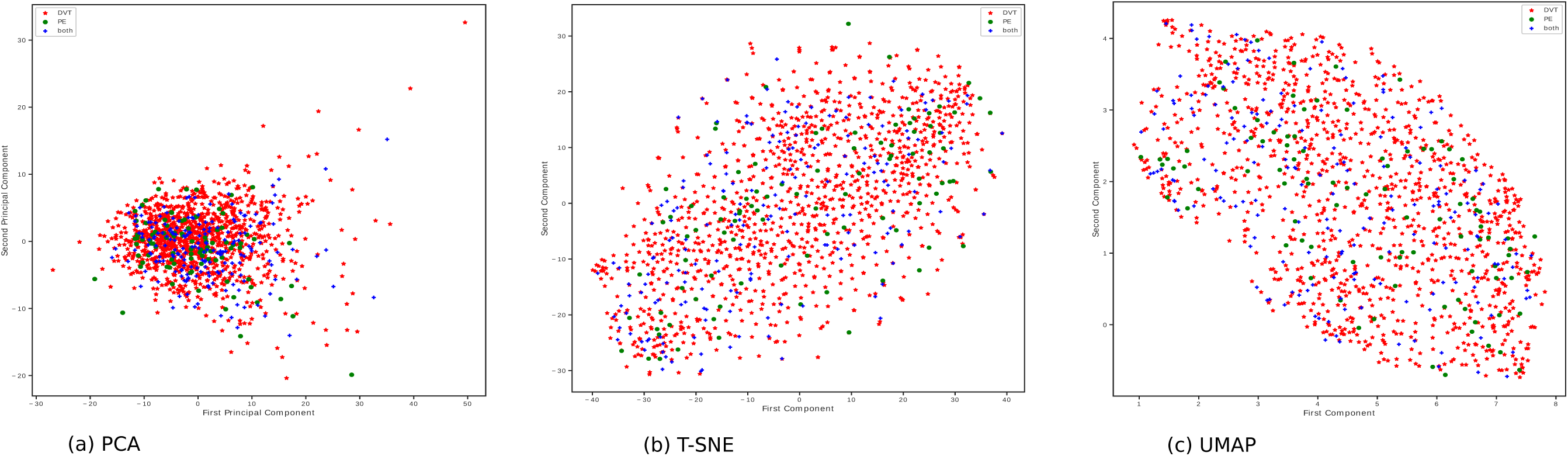
Graphical representation of the HPAs and biological MARTHA data projected on the first two principal components derived from standard principal components analysis (a), t-SNE (b) and UMAP (c) techniques.

### Artificial Neural Network for PE

As the accuracy/efficiency of any ANN strongly depend on the quality/homogeneity of the input data, we first applied the edited nearest neighbors algorithm [16] to perform under sampling of the majority class (DVT) and obtain a more homogeneous set of DVT patients, and further discarded the DVT+PE class to avoid adding noise in discriminating between PE and non PE patients. This strategy led to the selection of a subsample (referred thereafter to as the ANN sample 2) of 592 patients (497 DVT and 95 PE) whose proteomics/biological entered the ANN analysis.

A two hidden-layers ANN was then built from the ANN dataset with a training set of 576 patients (487 DVT and 89 PE) and a testing set of 16 patients (10 DVT and 6 PE). This allocation was chosen so that the number of PE cases used for training was sufficiently large. Because the training set presented with a strong imbalance with respect to the DVT/PE classes with ∼5 times more DVT than PE patients, the ANN was trained iteratively as described in the Materials and Methods section. By completion of the iterative algorithm, the final ANN obtained an area under the operative curve (AUC) of 0.89. Of more interest are the performances of the ANN in the testing set. Indeed, our ANN got F1-scores of 0.82 and 0.60 for the DVT and PE classes, respectively, and a global AUC of 0.79 in the testing set.

We then used the LIME algorithm to obtain a local linear approximate of the ANN predictions. In the testing set, the LIME prediction achieved an overal AUC of 0.77 instead of 0.79 for ANN. For each of the 16 patients in the testing set, we compared the individual predictions of their observed VTE event provided by the ANN and LIME methods (Table 2). In general, ANN and LIME predictions were rather consistent even if the ANN predictions seem to be more accurate in predicting DVT while LIME appears slightly more accurate in predicting PE. The average prediction in correctly classifying DVT patients was 0.872 by ANN compared to 0.748 by LIME. Note that one DVT patient (individual 10) was wrongly predicted to be PE by the ANN predictor, but not by the LIME predictor. Conversely, the average prediction in correctly classifying PE patients was 0.498 by ANN compared to 0.578 by LIME. Two PE patients (individuals 11 & 12) presented low predictions of being PE, using both ANN and LIME predictors.

We then assessed the correlation of the LIME predictor with the available biological phenotypes. No strong correlation was observed (Supplementary Table 3). However, the LIME predictor showed marginal positive correlation with fibrinogen (ρ = 0.12, p = 5.7 × 10^−3^) and factor VIII (ρ = 0.16, p = 0.013) plasma levels, and marginal negative correlation with prothombin time (ρ = −0.10, p = 0.029) and protein S (ρ =-0.10, p = 0.021) plasma levels. To go further into the biological interpretation of the LIME predictor, we sought to identify which proteins contribute the most to the definition of the LIME predictor. Figure 3 display the top 20 most contributing antibodies/proteins. Of note, 5 proteins tended to have substantial more importance than the remaining ones, among which three include proteins that had been selected because their gene expression (COX4I2, VCL, VWF) was found to be specifically enriched in endothelial cells [22].

**Table 3.**
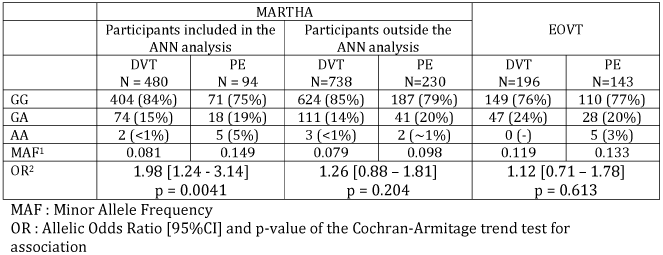
Association of rs1424597 with PE risk in the MARTHA and EOVT studies

**Figure 3.**
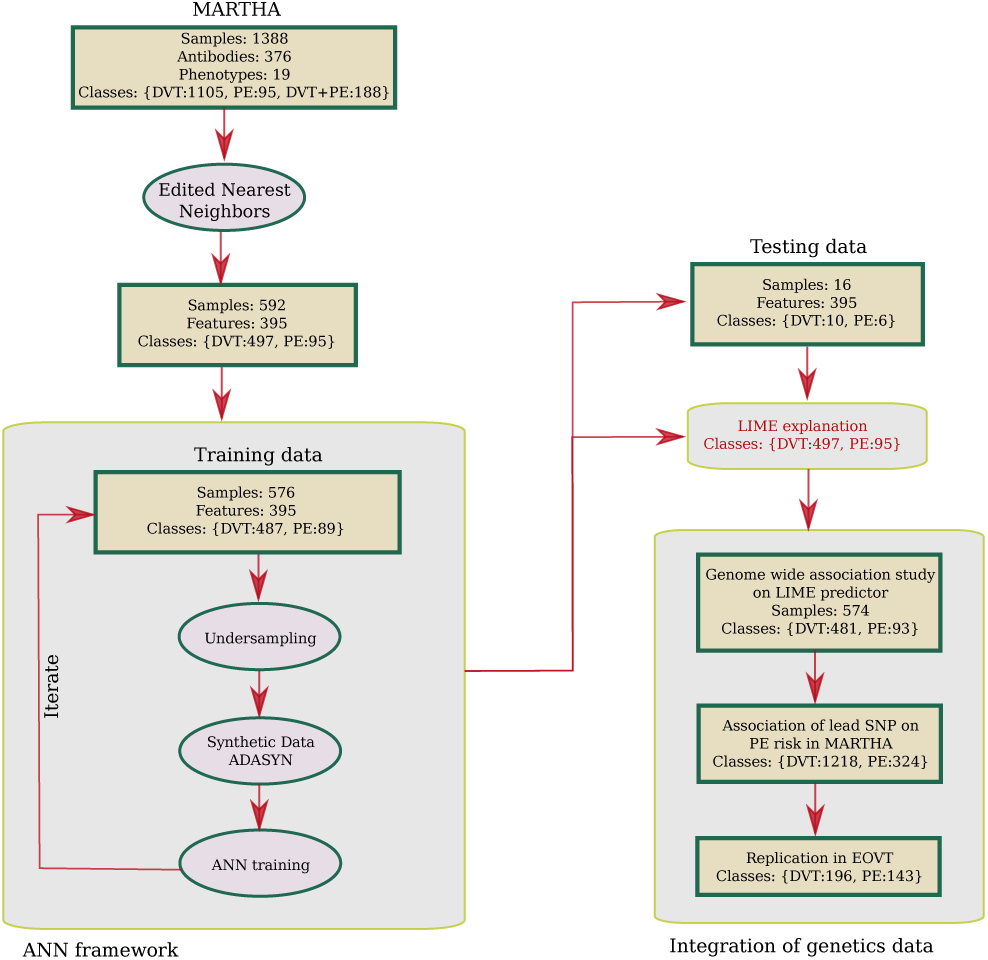
List of the top 20 antibodies contributing the most to the prediction model for PE

### Genetics of the LIME predictor

To get additional information about the biological mechanisms that could underly the linear LIME predictor, we conducted a GWAS on this predictor considered as a quantitative linear trait in a sample of 574 individuals of the ANN subsample with GWAS data. While no SNP reached genome-wide significance, we observed a peak of strong suggestive statistical association on chromosome 7 at the *PLXNA4* locus (Supplementary Figure1 – Supplementary Table 4). The sentinel SNP (p = 5.33 × 10^−7^) was rs1424597 whose minor A allele with frequency of 0.09 was associated with an increase of +0.169±0.034 in LIME predictor values. In this subsample, the rs1424597-A allele was slightly more frequent in patients with PE than in patients with DVT only (0.15 vs 0.08, p = 4.1 × 10^−3^) (Table 3). The association of rs1424597 with PE risk was then assessed in the remaining MARTHA samples (738 DVT and 230 PE (DVT+PE or isolated PE) patients) with available GWAS data and that had not been used for building our ANN model. In this subsample, we observed a trend for an higher frequency of the rs1424597-A allele in PE patients compared to non PE patients (0.10 vs 0.08), even if the association did not reach significance (p = 0.20).

We further investigated the association of rs1424597 with PE in the EOVT study composed of 143 PE patients 196 DVT patient. In EOVT, the rs1424597-A allele frequency was similar between EOVT patients with PE and with DVT (0.13 vs 0.12, p = 0.61) (Table 3). Because by design the EOVT study is enriched with early onset VTE patients, we assessed whether the discrepancy between MARTHA and EOVT results could be due to patient selection criteria (i.e according to age.) We thus split the EOVT samples according to the median of age of VTE onset, that was 37yrs. As shown in Table 4, the pattern of association of rs1424597 with PE slightly differed according to age. In EOVT patients younger than 37yrs, its allele frequency tend to be slighty lower in PE than in DVT patients (0.11s 0.13) while the inverse was observed in patients aged more than 37 yrs (0.15 vs0.11). Interestingly, the same observations hold in MARTHA when patients were stratified according to the same age threshold (Table 4). In the combined MARTHA and EOVT samples, the rs1424597-A allele was associated with an increased odds ratio (OR) for PE of 1.74 [1.27 – 2.38] (p = 5.42 × 10^−4^) in patients over 37 years of age while no association (OR = 0.96 [0.65 – 1.41], p = 0.848) was observed in younger patients. Similar ORs were obtained, OR = 1.73 [1.22 – 2.45] (p = 1.99 × 10^−3^) and OR = 1.08 [0.77 – 1.53] (p = 0.628), respectively, if a more standard age threshold of 40 yrs [23] had been used.

**Table 4.** Association of rs1424597 with PE risk according to age of onset of venous thrombosis

|  | Age of onset <37 |  |  |  | Age of onset ≥37 |  |  |  |
| --- | --- | --- | --- | --- | --- | --- | --- | --- |
|  | MARTHA |  | EOVT |  | MARTHA |  | EOVT |  |
|  | DVT<br>N=550 | DVT+PE/PE<br>N =147 | DVT<br>N=96 | PE<br>N=64 | DVT<br>N=668 | DVT+PE/PE<br>N =177 | DVT<br>N=100 | PE<br>N=79 |
| GG | 464 | 123 | 70 | 51 | 564 | 135 | 79 | 59 |
| GA | 83 | 23 | 26 | 12 | 102 | 36 | 21 | 16 |
| AA | 3 | 1 | 0 | 1 | 2 | 6 | 0 | 4 |
| MAF<sup>1</sup> | 0.081 | 0.085 | 0.135 | 0.109 | 0.079 | 0.136 | 0.105 | 0.152 |
| OR<sup>2</sup> | 1.056 [0.664 – 1.678]<br>p = 0.817 |  | 0.784 [0.392 – 1.566]<br>p = 0.470 |  | 1.820 [1.266 – 2.617]<br>p = 1.16 10<sup>-3</sup> |  | 1.53 [0.82 – 2.86]<br>p = 0.196 |  |

MAF : Minor Allele Frequency
OR : Allelic Odds Ratio [95%CI] and p-value of the Cochran-Armitage trend test for association

### Genetics of inconsistent LIME predictions

As shown in Table 2, our ANN/LIME models failed to correctly predict the true VTE outcome in four individuals from the testing set (individuals 10, 11, 12 and 15). First, it is worthy of note that these 4 individuals were all females. Second, the 3 female PE patients wrongly predicted to be DVT (individuals 11, 12 and 15) were all under oral contraceptives (OC) at the time of the PE event (age 45, 35 and 53, respectively), but not individual 10 incorrectly predicted to be PE. While we cannot rule out the possibility that our ANN/LIME models poorly behave in women under OC, we nevertheless sought to investigate whether discordant predictions could be due to genomic outlier individuals harboring very rare disease causing mutations that could make the global ANN/LIME predictions inaccurate, inline with the idea that the discrepancy between (deep learning derived) predicted and observed phenotypes could be a heritable trait [24]. Among these 4 individuals, only two (Individuals 11 and 15) have been sequenced for their whole genome. Sequence data of these two individuals were then scrutinized for candidate rare variants that could explain the VTE phenotype.

Individual 11 is a woman that experienced PE under oral contraceptives (OC) at age 45. Of note, her ten closest neighbors inferred from HPA data were all DVT patients which would likely explain why the derived ANN predicted her a DVT outcome instead of PE. She was not found to harbour any candidate variation in known VTE genes but presented in her genome with 61 very rare coding variants with strong predicted deleteriousness that could be good candidates responsible for the PE event.

Individual 15 is a woman that had experienced PE at age 53 also under OC. Nine out of 10 of her closest proteomics based neighbors were DVT patients which may also explain why this PE patient was uncorrectly predicted to be DVT. This patient was found to carry a very rare nonsynonymous variation (rs121918154; PROC:NM_000312:exon9:c.C814T:p.R272C) in the VTE-associated *PROC* gene. This variation has a minor allele frequency of 0.005% in public database (https://www.ncbi.nlm.nih.gov/snp/rs121918154), is predicted to be deleterious by several bioinformatic tools and have been previously reported in VTE patients with protein C deficiency [25,26]. This variation is located in the last exon of the gene and is predicted to alter splicing regulatory elements [27–29], which could lead to a deletion of a part of the peptidase S1 domain that is responsible for the clivage activity of the protein. Of note, this patient exhibited moderately low plasma Protein C levels, 63%, slightly lower than the 65% threshold adopted to declare moderate protein C deficiency [30].

## Discussion

This work is original in at least three main aspects. First, it is the largest plasma proteomic study with respect to pulmonary embolism in VTE patients. Second, it is to our knowledge the first attempt to deploy ANN methodologies on proteomic data with the aim at identifying new molecular thrombotic players. And finally, the integration of proteomic and genomics data identified *PLXNA4* as a new candidate gene for PE. This work started with the implementation of an ANN methodology on antibody based affinity proteomics data in relation to PE risk. This ANN was not developped as a tool to be used in clinic for predicting PE risk as 1/one is not 100% certain about the identity of the identified tagged proteins [31] (further experimental validation would be needed to assess this) and 2/ plasma protein levels determined with the antibody suspension bead array are not absolute but relative values depending on the current set of studied antibodies. Rather, our ANN based predictor for PE was aimed at serving as an intermediate surrogate biomarker that could generate new knowledge about the (genetics) mechanisms involved in PE. By conducting a GWAS on the derived PE predictor and capitalizing on two case-control samples totalling 467 patients with PE and 1414 patients with DVT, we observed that the *PLXNA4* rs1424597-A allele was associated with a ∼2-fold increased risk of PE in VTE patients aged more than ∼40yrs. *PLXNA*4 codes for Plexin A4, which is part of a receptor complex involved in signal transduction of sempahorin 3A signals linked to cytoskeletal rearrangement, inhibiting integrin adhesion [32,33]. It has a role in axone guidance in nervous system development, and genetic variants in PLXN4 have been linked to risk of Alzhemier disease [34,35]. Based on RNA seq data from HPA, FANTOM and GTEx datasets, *PLXNA4* is expressed at medium/high levels in central nervous system, adipose, breast and female reproductive tract tissues, and low levels in a broad range of other tissues (https://www.proteinatlas.org/ENSG00000221866-PLXNA4/tissue), indicating roles outside the nervous system. RNA seq data from blood cell populations show expression in plasma cytoid dendritic cells, NK cells and some T-cell populations, and research based on animal studies suggest a role in immunity and immune function. It has been shown to be a negative regulator of T cell activation [36]. Besides, Wen et al [37] found it to be highly expressed in myeloid cells, where PLXNA4 had an important function in stimulating TNF-alpha and IL-6 production in macrophages, where knock out mice were protected against letal dose LPS induced cytokine storms, suggesting it having a critical role in mediating pro-inflammatory cytokine production. The ligand of PLXNA4, SEMA3, has also been described with a role in endothelial cell function in an autocrine loop, promoting processes involved in vascular remodeling [38], and also in negatively regulating platelet aggregation [39]. While PLXNA4 thus has been described with a role in processes/pathways of relevance for thrombosis, little is known about *PLXNA4* in pulmonary embolism. In addition, we did not identify strong elements supporting a functional role of the intronic rs1424597 polymorphisms or of any other polymorphisms in strong linkage disequilibrium with it. Nevertheless, rs1424597 has recently been observed in the FinnGen study (http://r3.finngen.fi/) to be marginaly associated (p = 4.5 10^−3^) with pleural conditions that are inflammatory disorders of the lung. Consistent with this observation, we observed a positive correlation between the rs1424597-associated PE predictor and fibrinogen, a well known inflammatory marker. Additional *PLXNA4* polymorphisms have also been reported to demonstrate strong statistical evidence for association with various lung function markers [40,41]. Besides, our group have previously suggested that *PLXNA4* polymorphisms could interact with polymorphisms at *UQCC1* to modulate the risk of VTE in the general population [42], *UQCC1* being a locus that have also be shown to be involved in lung function [43]. Altogether, these observations strongly support for a role of *PLXNA4* in lung function and its precise role in the etiology of pulmonary embolism deserve further investigation. Which polymorphisms could be truly responsible for the observed association with PE risk also merits further works as the rs1424597 is likely tagging for functional variant(s)/haplotypes yet to be characterized. Finally, further studies would be needed to investigate whether the previously suggested *PLXNA4* x *UQCC1* interaction on the risk of VTE (combining both DVT and PE) could be more specific to patients with PE.

In addition to searching for common polymorphisms that could associate with our ANN based predictor and with PE risk, we also looked for rare variants that could explain the discrepancy between predicted and observed VTE outcome in our testing set. Two out of four patients with discordant predictions in the testing set have been sequenced for their whole genome. Both were females patients that experienced PE under OC. In one of them, we were able to identify a rare VTE causing mutation in *PROC*. It is not our intention to conclude to any general rule about the relevance of searching of rare variants responsible for any discordancy between ANN predictions and observed outcomes. Especially as we observed that the three PE patients wrongly predicted to be DVT were women who developed PE under OC. These observations could suggest that our plasma proteomics ANN derived predictions may not be valid in such subgroups of VTE patients and highlight the challenge to identify general prediction models for complex diseases. Several additional limitations must be addressed. First, no plasma antibody targeting PLXNA4 was available when the screening phase of this work was initiated preventing us from validating further its association with PE. Second, no proteomic data was available in the EOVT study to formally replicate the association of our ANN and LIME predictors with PE risk. Third, our GWAS analysis on the ANN derivedpredictor was performed only in 574 samples which has likely hampered our power to identify genome-wide significant SNPs. We may have then missed additional polymorphisms that could be truly associated with the predictor and could have then help us to better dissentangle its underlying molecular biology. Finally, the moderate sample size of the EOVT study has also likely hampered our power for statistically replicating the association of the lead *PLXNA4* polymorphism with PE. In addition, no information was available in the EOVT study to distinguish isolated PE From DVT+PE which prevented us from further testing whether the association of *PLXNA4* with PE risk was mainly restricted to isolated PE as suggested from the MARTHA results. However, the very consistent pattern of association observed according to age strata between the two studies is a strong argument in favor of *PLXNA4* as a new candidate in PE biology. In conclusion, by implementing an original artificial neural network methodology integrating plasma proteomics and genetic data, we identified *PLXNA4* as a new candidate susceptibility gene for PE in VTE patients whose precise role in PE etiology deserve further investigations

## Materials and Methods

### Ethical approval

Each individual study on which the work is based was approved by its institutional ethics committee and informed written consent was obtained in accordance with the Declaration of Helsinki. Ethics approval were obtained from the “Departement santé de la direction générale de la recherche et de l’innovation du ministère” (Projects DC: 2008-880 & 09.576) and from the institutional ethics committees of the Kremlin-Bicetre Hospital.

### MARTHA study

The MARTHA population is composed of VTE patients recruited from the Thrombophilia center of La Timone hospital (Marseille, France) and free of any chronic conditions and of any well characterized genetic risk factors including antithrombin, protein C or protein S deficiency, homozygosity for FV Leiden or Factor II 20210A, and lupus anticoagulant. Detailed description of the MARTHA population has been provided elsewhere [14,44].

#### MARTHA proteomics substudy

A sample of 1,388 MARTHA patients with available plasma samples were profiled for targeted plasma proteomic investigations as described below.

#### MARTHA genetic substudy

From the whole MARTHA population, 1592 patients with DNA available were genotyped with high-throughput genotyping arrays (see below).

### Plasma proteomic profiling

#### Generation of antibody suspension bead array (SBA)

The multiplex antibody suspension bead array (SBA) was created by covalent coupling of 339 Human Protein Atlas (HPA) antibodies, 13 from commercial providers and 25 monoclonal BSI antibodies (BioSystems International Kft) targeting 234 unique candidate proteins (Supplemental Table 2). Antibodies were individually coupled to carboxylated magnetic beads (MagPlex-C, Luminex Corp.) generating up to 384 different bead identities (IDs), essentially according to methods previously described [45,46]. The final multiplexed suspension bead array was prepared by combining all 384 antibody coupled beads into a single SBA stock with a concentration of approximately 25-40 beads of each antibody bead ID/ul.

#### Plasma labelling and protein profiling assay

Plasma samples were diluted 1:10 in filtered 1xPBS and labelled with biotin (NHS-PEG4-Biotin, Thermo Scientific) for 2h at 4°C. The labelling process was terminated by the addition of 12,5ul of 0.5M HCl pH:8.0 to each sample for 20 min and consecutively storage at −20°C until usage [45]. Labelled plasma samples were diluted 1:50 in PVX casein buffer + 10% (v/v) rabbit IgG (0.1% casein, 0.5% polyvinyl alcohol, 0.8% polyvinylpyrrolidone, prepared in 1xPBS). Diluted samples were heat-induced to achieve epitope retrieval for 30 minutes at 56°C. Five microliters of the SBA were mixed with 45ul of heat-treated samples for 16-18 hours, at RT and constant shake. Unbound complexes were removed by 2 consecutive washes with PBS-T and antibody-bound complexes were cross-linked by resuspending the beads in 0.4% PFA-PBS for 10 min. R-phycoerythrin-conjugated streptavidin (1:750, PBS-T; Invitrogen) was added to all samples for 30 min followed by 2 times washes. Relative amount of each protein complex was expressed as median of fluorescence intensity (MFI) by read out on a FlexMAP3D.

### The Early Onset Venous Thrombosis (EOVT) study

This study is composed of 339 VTE patients with documented idiopathic isolated PE or DVT selected according to the same criteria as the MARTHA participants, with the exception that the age of VTE onset was below 50 yrs. Detailed description can be found in [44,47]

### Deep-learning framework for identifying a molecular predictor of PE risk

#### Step 1: Normalization

First, all HPA variables were normalized and scaled to have 0 mean et 1 variance to avoid major artificial influence of variables with large range of variations.

#### Step2: Edited nearest neighbors

As our aim was to identify new molecular markers associated with PE, we hypothesized that conducting our discovery phase on isolated PE, an expected less heterogeneous class of VTE patients than the class of patients with both DVT and PE, will increase our chance to identify novel relevant molecular players. As a consequence, we decided to built our ANN model only on patients with isolated PE (N = 95) or with DVT (N = 1105). However, due to the imbalance nature of this dataset with ∼10 more samples in the DVT class than in the PE class, we applied the edited nearest neighbors (ENN) algorithm, an under sampling method usually used in the field of pattern recognition or classification in presence of unbalanced samples [16]. This method relies on under sampling unit of analysis, in our case individuals, from the majority class by removing the most heteregenous units. It consists in computing the Euclidian distance between each pair of individuals from their proteomics data and to remove samples whose clinical phenotype (here DVT) is not consistent with that of his/her k nearest neighbors (k=3 in this work). This led us to the selection of the so called ANN dataset composed of N = 497 DVT and N = 95 PE patients for building our ANN model.

#### Step3: Derivation of an ANN model for PE prediction

To build our ANN model, the ANN dataset was divided into a training set composed of 576 patients (487 DVT and 89 PE) and a testing set of 16 patients (10 DVT and 6 PE), the latter being used for testing the accuracy of the ANN model derived from the former. This allocation was chosen so that the number of PE cases used for training was sufficiently large.

Because the application of a standard ANN methodology to our training set would lead to instable network for predicting PE due to the imbalance nature of the input data with ∼5 times more DVT than PE patients, an interative ANN framework was adopted:

At each iteration *i*,

- A random sample of 30 PE patients and 100 DVT patients is selected from the training set and a sample of 70 synthetic PE samples are generated using the ADASYN algorithm [48]. ADASYN is an adaptive synthetic data generation method where new samples are generated based on the weighted distribution for minority class samples with two main advantages, resolving data imbalance and forcing classifiers to be more sensitive to the minority class. This strategy led to a balanced dataset D*i* of 100 PE and 100 DVT (synthetic) patients on which a ANN is built.
- Using the Di dataset further splitted randomly into 90%/10% training/testing subsamples, a two hidden-layers feed forward neural networks was implemented. The first hidden layer has 395 neurons corresponding to the number of input (proteomic & biological) variables, the second layer 128 while the output layer consisted in 2 neurons, representing the DVT and PE classes respectively. The number of neurons were selected by trial and error approach under the constraint that the number of neurons shall be smaller than the number of input variables and higher than the number of output classes

The Rectified Linear Unit (ReLU) function [49]was used to activate hidden layers while the softmax activation function [50]was used to generate class probabilities in the output layer.

After fixing the number of nodes, layer and activation function, the process of training the neural network can start. Starting from random weights, forward propagation is used to generate the output of all nodes at all layers while moving from the input to the output layers. The generated final output is compared to the observed class phenotype and an error is calculated using the cross-entropy function [51]. Iteratively, this error was then backpropagated using a gradient descent algorithm [52] (with learning of 0.01 and batch size of 32) to update weights according to their contribution to the error. In order to reduce over-fitting and obtain the best performing model, the callback feature proposed by the Keras open-source library (https://keras.io/) that was employed for developping this ANN framework was used.

#### Step4: Local Interpretable Model-Agnostic Explanations (LIME)

As a neural network is often considered as a black box without telling much about which, and how, input variables contribute the most to the predictive model, the LIME methodology [17] was applied to the final ANN model obtained at Step 3 in order to inform about which input variables (i.e plasma proteoin levels) contribute top PE risk prediction and what are the relative weights using a linear approximation of the ANN model.

### Genome Wide Genotyping

As previously described [15,44], both MARTHA and EOVT participants have been genotyped with high-density genotyping Illumina arrays and imputed for single nucleotide polymorphisms (SNPs) from the 1000G Phase I Integrated Release Version 2 Haplotypes using MACH (v1.0.18.c) and Minimac (release 2011-10-27) imputation software.

### Genome-Wide Association analysis (GWAS)

Imputed SNPs with imputation quality *r*^2^ greater than 0.5 and with minor allele frequency (MAF) greater than 0.01. were tested for association with the LIME predictor derived in 574 MARTHA participants. Associations with statistical p-value < 5 × 10^−8^ were considered as genome-wide significant.

### Genetic Association Analysis with PE risk

The candidate SNP identified from the GWAS on the LIME predictor was tested for its association with PE risk, both in MARTHA and EOVT participants. For this, we employed the Cochran-Armitage trend for association applied to the best guessed genotypes inferred from the imputed allele dosage at the SNP of interest. Meta-analysis of the association results observed in MARTHA and EOVT was conducted using a fixed-effects model based on the inverse-variance weighting and heterogeneity of association between the two studies was assessed by the Cochran-Mantel-Haenszel test statisticsn-Mantel-Haenszel test statistic [53]

### Whole Genome Sequencing

From the whole MARTHA study, 200 patients had been selected for whole genome sequencing. These patients were selected to have experienced unprovoked VTE. Besides, these patients should have family history of VTE or multiple unprovoked VTE events, such clinical patterns being compatible with the existence of an underlying VTE causing genetic defect. Genomic DNA was extracted from peripheral blood, using the BioRobot EZ1 workstation. The DNA concentration was determined using the Qubit assay kit (Thermofisher). Whole genome sequencing was performed at the Centre National de Recherche en Génomique Humaine (CNRGH, Institut de Biologie François Jacob, Evry, FRANCE). After a complete quality control, 1µg of genomic DNA was used for each sample to prepare a library for whole genome sequencing, using the Illumina TruSeq DNA PCR-Free Library Preparation Kit, according to the manufacturer’s instructions. After normalisation and quality control, qualified libraries were sequenced on a HiSeqX5 instrument from Illumina (Illumina Inc., CA, USA) using a paired-end 150 bp reads strategy. One lane of HiSeqX5 flow cell was used per sample specific library in order to reach an average sequencing depth of 30x for each sequenced individual. Sequence quality parameters have been assessed throughout the sequencing run and standard bioinformatics analysis of sequencing data was based on the Illumina pipeline to generate FASTQ file for each sample. FastQ sequences were aligned on human genome hg37 using the BWA-mem program [54]. Variant calling was performed using the GATK HaplotypeCaller (GenomeAnalysisTK-v3.3-0, https://software.broadinstitute.org/gatk/documentation/article.php?id=4148). Single-sample gVCFs files were then aggregated using GATK CombineGVCFs and joint genotyping calling performed by GATK GenotypeGVCFs. Recalibration was then conducted on the whole gVCF following GATK guidelines. Following GATK VQSR, we retained single nucleotide variants in the 99.5% tranche sensitivity threshold and indels in the 99% tranche sensitivity threshold for further analysis and annotated them using Annovar [55].

As a strategy to identify candidate variants that could explain the VTE phenotype in individuals with discordant class prediction, we first prioritized variants that were likely functional (stop loss/stop gain, frameshift, non-synonymous and splicing variants), located in known VTE associated genes (ABO, ARID4A, C4BPB, EIF5A, F2, F3, F5, F8, F9, F13A1, FGG, GRK5, MPHOSPH9, MAST2, NUGCC, OSMR, PLAT, PLCG2, PLEK1, PROC, PROS1, SCARA5, SERPINC1, SLC44A2, STAB2, STX10, STXBP5, THBD, TSPAN15, VWF) [56–58], that have not been reported or at a low frequency (<1‰) in public genomic data repositories (dbSNP, GnomAD) and that was present in only one of the 200 sequenced patients. If no candidate variants was identified in known VTE genes, we extended our search to whole coding genes and also took into account the predicted deleteriousness of selected candidates using *in silico* tools such as SIFT, PolyPhen and CADD-v1.2 [59] to further reduce the number of candidates.

## Data Availability

Data are available upon specific request to corresponding authors.

## Acknowledgments

Mi.R; O.S. and Ma.R and the production of the MARTHA genomics data were financially supported by the GENMED Laboratory of Excellence on Medical Genomics [ANR-10-LABX-0013], a research program managed by the National Research Agency (ANR) as part of the French Investment for the Future. This work benefited from the financial support from the "EPIDEMIOM-VTE" Senior Chair from the Initiative of Excellence of the University of Bordeaux. Bioinformatics and statistical analyses benefit from the CBiB computing centre of the University of Bordeaux. The proteomics screening was financed by a grant from Stockholm County Council (SLL 2017-0842) and from Familjen Erling Perssons Foundation.

## Supporting information captions

**Supplementary Figure 1** Manhattan plot describing the results of the genome wide association study on the LIME predictor

**Supplementary Table 1** List of biological phenotypes that have been measured in MARTHA patients

**Supplementary Table 2** List of Human Protein Atlas antibodies measured in MARTHA patients

**Supplementary Table 3** Correlation between LIME PE predictor and biological traits available in MARTHA participants used for building the ANN

**Supplementary Table 4** Main statistical associations (p< 1 × 10^−5^) observed in the GWAS on LIME predictor

**Supplementary Table 5** Rare coding variants identified by whole genome sequencing in individual 11.

## Notes

### Competing Interest Statement

The authors have declared no competing interest.

### Funding Statement

Mi.R, O.S, Ma.R, and the production of the MARTHA genomics data were financially supported by the GENMED Laboratory of Excellence on Medical Genomics [ANR-10-LABX-0013], a research program managed by the National Research Agency (ANR) as part of the French Investment for the Future. This work benefited from the financial support from the EPIDEMIOM-VTE Senior Chair from the Initiative of Excellence of the University of Bordeaux. Bioinformatics and statistical analyses benefit from the CBiB computing centre of the University of Bordeaux. The proteomics screening was financed by a grant from Stockholm County Council (SLL 2017-0842) and from Familjen Erling Perssons Foundation.

### Author Declarations

Each individual study on which the work is based was approved by its institutional ethics committee and informed written consent was obtained in accordance with the Declaration of Helsinki. Ethics approval were obtained from the Departement santé de la direction générale de la recherche et de l’innovation du ministère (Projects DC: 2008-880 & 09.576) and from the institutional ethics committees of the Kremlin-Bicetre Hospital.

