## Supplementary material for "An artificial neural network approach integrating plasma proteomics and genetic data identifies *PLXNA4* as a new susceptibility locus for pulmonary embolism": Supplementary Table S3.docx

Supplementary Table S3: Correlation between LIME PE predictor and biological traits available in MARTHA participants used for building the ANN

| Trait | ρ | P | N |
| --- | --- | --- | --- |
| Mean corpuscular volume | -0.042 | 0.314 | 574 |
| Mean platelet volume | 0.014 | 0.724 | 574 |
| Platelets count | 0.029 | 0.501 | 525 |
| Red blood cells count | 0.044 | 0.282 | 574 |
| White blood cells | 0.051 | 0.225 | 574 |
| Basophil | -0.004 | 0.932 | 574 |
| Eosinophil | 0.049 | 0.244 | 574 |
| Lymphocytes | -0.013 | 0.762 | 574 |
| Monocytes | 0.080 | 0.054 | 574 |
| Neutrophil | 0.075 | 0.073 | 574 |
| Plasminogene Activator Inhibitor-1 activity | -0.017 | 0.689 | 574 |
| Fibrinogen | 0.115 | 5.67 10^-3^ | 574 |
| Factor VIII | 0.159 | 0.013 | 249 |
| Factor XI | -0.015 | 0.708 | 574 |
| von Willebrand Factor | 0.047 | 0.299 | 479 |
| Prothrombin Time | -0.095 | 0.029 | 529 |
| Antithrombin | 0.045 | 0.314 | 493 |
| Protein C | -0.068 | 0.126 | 500 |
| Protein S | -0.103 | 0.021 | 496 |
