## Supplementary material for "An artificial neural network approach integrating plasma proteomics and genetic data identifies *PLXNA4* as a new susceptibility locus for pulmonary embolism": Supplementary TableS1.docx

**Supplementary Table 1 : List of biological phenotypes that have been measured in MARTHA patients**

Factor VIII, Factor XI, von Willebrand Factor, prothrombin time, antithrombin, protein C, protein S, PAI-1, platelets count, red blood cells count, mean platelet volume, mean corpuscular volume, white blood cells count, neutrophils, eosinophil, basophil, lymphocytes, monocytes, fibrinogen,
