## Supplementary figures and images for "An artificial neural network approach integrating plasma proteomics and genetic data identifies *PLXNA4* as a new susceptibility locus for pulmonary embolism"

### Supplementary Figure 1.png

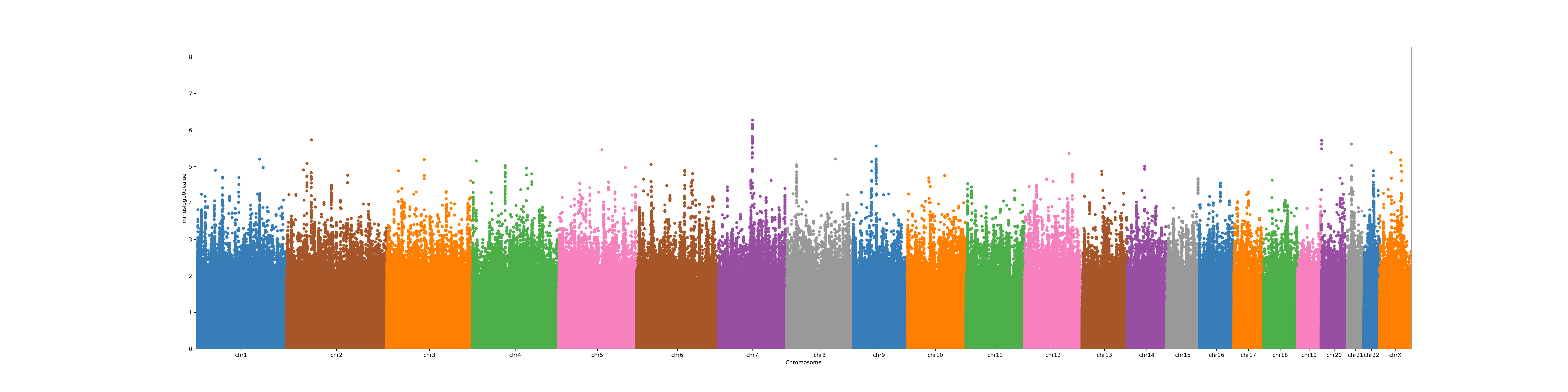
